# Patterns of Multimorbidity

**DOI:** 10.1101/2021.05.13.21256888

**Authors:** Kien Wei Siah, Chi Heem Wong, Jerry Gupta, Andrew W. Lo

## Abstract

**Background:** With multimorbidity becoming the norm rather than the exception, the management of multiple chronic diseases is a major challenge facing healthcare systems worldwide.

**Methods:** Using a large, nationally representative database of electronic medical records from the United Kingdom spanning the years 2005 to 2016 and consisting over 4.5 million patients, we apply statistical methods and network analysis to identify comorbid pairs and triads of diseases and identify clusters of chronic conditions across different demographic groups. Unlike many previous studies, which generally adopt cross-sectional designs, we examine temporal changes in the patterns of multimorbidity. In addition, we perform survival analysis to examine the impact of multimorbidity on mortality.

**Results:** The proportion of the population with multimorbidity has increased by approximately 2.5 percentage points over the last decade, with more than 17% having at least two chronic morbidities. We find that the prevalence and the severity of multimorbidity increase progressively with age. Stratifying by socioeconomic status, we find that people living in more deprived areas are more likely to be multimorbid compared to those living in more affluent areas at all ages. The same trend holds consistently for all years in our data. In addition to a number of strongly associated comorbid pairs (e.g., cardiac-vascular and cardiac-metabolic disorders), we identify three principal clusters: a respiratory cluster, a cardiovascular cluster, and a mixed cardiovascular-renal-metabolic cluster. These are supported by established pathophysiological mechanisms and shared risk factors, and are largely consistent with existing studies in the medical literature.

**Conclusions:** In this paper, we use data-driven methods to characterize multimorbidity patterns in different demographic groups and their evolution over the past decade. Our findings contribute to the better understanding of the epidemiology of multimorbidity that is needed to develop more effective primary care for multimorbid patients.

## 1 Introduction

Multimorbidity, defined as the coexistence of two or more chronic medical conditions in an individual patient^1^, is a growing public health concern for healthcare systems worldwide. It has been found to be associated with adverse health outcomes, including a higher risk of mortality, a lower quality of life, increased utilization of healthcare, and correspondingly higher healthcare costs.^2–14^ It is most prevalent in the elderly population, as organs gradually lose full function with the aging process.^8,15–17^ With an increasing life expectancy and an aging population, the number of people with multiple health conditions is set to rise, as is public expenditure on long-term medical care. Unfortunately, current healthcare systems are largely designed to treat single diseases, resulting in the need to use multiple services to manage multimorbidity.^11,18–20^ Due to poor coordination and integration in medical care, causing a lack of continuity in treatment, disorders not designated as the primary condition are often undertreated.^21^

In order to align primary care more closely to the needs of patients suffering from multiple health conditions, a better understanding of the epidemiology of multimorbidity in the general population is necessary. However, previous studies on multimorbidity have mostly been limited in focus to elderly patients, due to its high prevalence in that population.^2,19,20,22– 25^ Since this data is typically collected through self-administered questionnaires and research interviews, it may be subject to self-reporting bias. Analyses based on small sample sizes from selected populations are known not to generalize well. Furthermore, many studies employ only a narrow range of methods to study multimorbidity patterns.

In this paper, we aim to characterize multimorbidity patterns not only in older patients, but also across groups with different demographic and socioeconomic statuses, using a large, nationally representative primary care electronic medical records database. We apply standard statistical methods to identify common comorbid pairs and triads of diseases, and use network analysis algorithms to identify clusters of chronic conditions. Unlike many previous studies, which adopted a cross-sectional design (i.e., analyzing data at a single point in time), we examine temporal changes in the patterns of multimorbidity across a decade of patient data. In addition, we analyze the impact of multimorbidity on mortality using survival models. In the discussion, we compare our findings with related works in literature.

## 2 Methods

### a Data

We use anonymized electronic medical records from The Health Improvement Network (THIN)^26^ database for our analysis. The database contains longitudinal patient data collected at primary care clinics throughout the UK, covering approximately 6% of the UK population. We extract demographic information (e.g., date of birth, sex, geographical location, and socioeconomic group), baseline vitals (e.g., smoking and alcohol status), and medical history (e.g., medical condition and date of diagnosis) from patient records between 2005 and 2016. We categorize the subjects into seven mutually exclusive age groups based on Medical Subject Headings (MeSH) definitions (see Appendix A).^27^

Diagnoses are recorded in the THIN database using Read Codes, a coded thesaurus of clinical terms used by the UK National Health Service since 1985.^28^ There is no standard method for the selection and definition of morbidities in the literature. After consulting with medical officers and Life & Health (L&H) actuaries at Swiss Re, we identify chronic conditions in the records, that is, diseases that are either permanent, caused by nonreversible pathological alterations, or require long periods of rehabilitation and care,^19,29^ and map them to a list of 46 higher level morbidities. Furthermore, we classify the morbidities into 14 System Organ Classes (SOCs) as defined in the Medical Dictionary for Regulatory Activities (MedDRA) dictionary. (See Appendix A for list of morbidities and classifications.) As in similar studies, we define multimorbidity as the presence of at least 2 of the 46 morbidities in a patient.

### b Statistical analysis

We examine the distribution of multimorbidity in relation to age and socioeconomic status, as done in Barnett et al.^11^ However, we use the Index of Multiple Deprivation (IMD) as a proxy for socioeconomic status. The IMD is a widely used measure of relative deprivation or poverty of wards and districts in the UK. It is computed using census data as a weighted index of deprivation in seven domains, including income, employment, education, health, crime, barriers to housing and services, and living environment.^30^ (IMD data was available only for a subset of the patients. See Appendix B for the sample sizes used in this analysis.) We note that the same approach, defining socioeconomic status by the area of residence, has been used in previous studies.^11,31^

For each age group, we also compute the observed prevalence for all individual, pairs, and triplets of morbidities. By the assumptions of probability theory, we expect diseases that are independent to co-occur at a rate close to the product of the observed prevalence of each individual constituent disease (i.e., the expected prevalence). Therefore, by comparing the ratio of the observed prevalence versus the expected prevalence (i.e., the lift), we can identify pairs and triads of diseases that occur together more frequently than expected by chance, possibly driven by an underlying pathophysiological mechanism. As a second metric, we estimate the odds ratio using logistic regression models to determine the association between each pair of diseases, both without adjustment and adjusted by age, sex, and all other diseases.

Next, we construct multimorbidity networks to study the natural clustering of diseases in the dataset. We consider diseases as nodes with sizes proportional to their observed prevalence. For each pair of diseases, we connect their nodes with an undirected edge weighted by the estimated lift, a measure of the strength of the association between the comorbid pair. This creates a dense network where each node is linked to almost every other node. This density, however, makes visualization and inference difficult. As a pre-processing step for subsequent analysis, we extract the main graph structure by removing edges from the adjacency matrix that are peripheral and relatively unimportant. We prune the edges between nodes that have joint prevalence below the 90^th^ percentile, and keep only the edges that have a lift above 2.0, i.e., those edges between pairs that co-occur two times more frequently than expected by chance. Similar thresholds have been used in related studies.^2,32–35^

We compute measures of centrality to identify the most important vertices in the multimorbidity network. In particular, for each node, we compute the degree centrality, which is defined as the number of links incident on a node, a direct measure of the connectivity of a node. In this context, a disease with high degree centrality is important because it often co-occurs with a large number of pathologies. We also estimate the eigenvector centrality, a measure of the transitive influence of nodes. To calculate the eigencentrality, each node is assigned a score that is proportional to the sum of the scores of all of its neighbors. Nodes with high eigencentrality either have many connections, or are connected to important neighbors. In addition, we compute the graph clustering coefficient (also known as the transitivity) as a quantitative measure of the network’s tendency to aggregate in smaller subgroups. To identify any clusters embedded in the multimorbidity networks, we apply a community detection algorithm based on modularity maximization^36– 38^ to partition nodes into groups that have dense intra-group connections and sparse inter-group connections.

To gain insight into temporal disease associations, we construct directed multimorbidity networks. We extract from each patient’s medical history a sequence of diseases ordered by the time of diagnosis. Using these trajectories, we can derive the probability of any given disease conditional on some prior diagnosis, i.e., Prob(Disease B given Disease A). We use these probabilities as weights of the directed edges in the network. As before, we prune the network based on node prevalence and edge weights. Since these connections are directed, we can compute the in-degree and out-degree centralities, defined as the number of edges directed to the node, and the number of edges directed from the node to others, respectively. A node with a high in-degree centrality is often diagnosed following other diseases; a node with a high out-degree centrality often leads to subsequent diagnoses in other diseases. These metrics are useful for understanding disease progression, and any causal or contributory relationships between diseases.

Finally, we examine the impact of multimorbidity on mortality by performing survival analysis on the dataset. We use the five-year overall survival as the primary outcome variable, and consider in our models a range of features, including demographic group, baseline vitals, baseline medical history, the severity of multimorbidity as quantified by the number of co-occurring chronic conditions, and the presence of any of the top ten most prevalent pairs and triplets of morbidities as observed in the aged and elderly age groups. We exclude those subjects aged 65 or less from this part of the analysis, as younger age groups have five-year overall mortality rates close to zero.

We explore three standard methods used in survival modeling, the Cox proportional hazards model^39^, the regularized Cox model, and the accelerated failure time model, and additionally, we apply a nonlinear and non-parametric neural network survival model.^40^ For model estimation and validation, we randomly split the original dataset into two disjoint sets, a training set that comprises 70% of the data, and a testing set that comprises the remaining 30%. We use the training set to estimate our models, and keep the testing set as an out-of-sample dataset for performance validation. We use the concordance index (C-index) as the metric for model performance. This metric is commonly used in survival analysis to evaluate its predictive power.^41^ It is a measure of the concordance between orderings of observed survival times and the predicted times or risks. (A C-index of 0.5 corresponds to a random model, while a value of 1.0 corresponds to a perfect model.) We use cross-validation to tune the hyperparameters of the models.

In addition to discriminative power, we assess the calibration of our models by comparing the actual and the predicted survival probabilities at 36, 48, and 60 months of overall survival. For each time cutoff, we divide the test set into quintiles based on the predicted risk scores. We then compute the average predicted score and the true survival probability observed in each of the quintiles. Lastly, we create calibration plots by plotting the observed probabilities against the predicted probabilities. In the ideal case, the points should lie as close as possible to the diagonal line, which represents perfect calibration.

## 3 Results

We summarize the demographic statistics of the study population in Table 1. On average, the dataset consists of approximately 4.6 million patients each year, with an even mix of both sexes in all years. Most of the patient records were collected in England, which makes up the largest part of the population of the United Kingdom. However, the distribution in geographical location has evolved over the years, shifting towards other regions in the country. Over 60% of the patients are in the Adult (19 to 45 years old) and Middle-Aged (45 to 65 years old) age groups, as defined by the MeSH classification (see Appendix A). Approximately 15% are over 65 years old.

**Table 1.** Demographics of the dataset between 2005 and 2016.

| Proportion (%) | 2005 | 2006 | 2007 | 2008 | 2009 | 2010 | 2011 | 2012 | 2013 | 2014 | 2015 | 2016 |
| --- | --- | --- | --- | --- | --- | --- | --- | --- | --- | --- | --- | --- |
| *<u>Sex</u>* |  |  |  |  |  |  |  |  |  |  |  |  |
| Male | 49.9 | 49.9 | 49.9 | 49.9 | 49.8 | 49.8 | 49.7 | 49.7 | 49.6 | 49.6 | 49.6 | 49.6 |
| Female | 50.1 | 50.1 | 50.1 | 50.1 | 50.2 | 50.2 | 50.3 | 50.3 | 50.4 | 50.4 | 50.4 | 50.4 |
| *<u>Country</u>* |  |  |  |  |  |  |  |  |  |  |  |  |
| England | 70.1 | 69.8 | 68.9 | 68.7 | 67.9 | 67.2 | 66.7 | 65.9 | 63.3 | 59.7 | 51.1 | 45.1 |
| Northern Ireland | 4.3 | 4.4 | 4.6 | 4.4 | 4.5 | 4.6 | 4.7 | 4.7 | 5.1 | 5.5 | 6.8 | 7.6 |
| Scotland | 15.6 | 15.6 | 16.0 | 16.3 | 16.7 | 16.9 | 17.0 | 17.4 | 18.7 | 20.5 | 24.9 | 27.8 |
| Wales | 9.9 | 10.1 | 10.5 | 10.7 | 11.0 | 11.4 | 11.6 | 12.0 | 12.9 | 14.2 | 17.2 | 19.4 |
| *<u>Age Group</u>* |  |  |  |  |  |  |  |  |  |  |  |  |
| Infant | 0.9 | 0.9 | 0.9 | 0.9 | 1.0 | 0.9 | 1.0 | 1.0 | 1.0 | 0.9 | 0.9 | 0.9 |
| Child | 12.5 | 12.5 | 12.5 | 12.5 | 12.6 | 12.6 | 12.7 | 12.8 | 12.8 | 13.0 | 13.0 | 13.1 |
| Adolescent | 6.6 | 6.6 | 6.6 | 6.6 | 6.6 | 6.6 | 6.7 | 6.7 | 6.7 | 6.8 | 6.8 | 6.8 |
| Adult | 35.6 | 35.4 | 35.2 | 34.9 | 34.7 | 34.3 | 34.0 | 33.8 | 33.3 | 33.0 | 32.9 | 33.1 |
| Middle-Aged | 27.2 | 27.4 | 27.6 | 27.7 | 27.7 | 27.9 | 27.8 | 27.6 | 27.6 | 27.6 | 27.7 | 27.7 |
| Aged | 12.5 | 12.4 | 12.5 | 12.6 | 12.7 | 12.8 | 13.1 | 13.4 | 13.7 | 13.8 | 13.9 | 13.7 |
| Elderly | 4.7 | 4.7 | 4.7 | 4.7 | 4.7 | 4.8 | 4.9 | 4.9 | 4.9 | 4.9 | 4.8 | 4.8 |
| *<u>Multimorbidity</u>* |  |  |  |  |  |  |  |  |  |  |  |  |
| 0 | 63.6 | 62.9 | 62.4 | 62.0 | 61.7 | 61.4 | 61.1 | 60.9 | 60.5 | 60.3 | 60.2 | 60.4 |
| 1 | 21.8 | 22.0 | 22.1 | 22.3 | 22.3 | 22.4 | 22.4 | 22.4 | 22.5 | 22.5 | 22.5 | 22.4 |
| 2 | 7.9 | 8.1 | 8.3 | 8.4 | 8.5 | 8.6 | 8.7 | 8.8 | 8.9 | 8.9 | 8.9 | 8.9 |
| 3 | 3.3 | 3.5 | 3.6 | 3.6 | 3.7 | 3.8 | 3.8 | 3.9 | 4.0 | 4.0 | 4.1 | 4.0 |
| 4 | 1.7 | 1.8 | 1.8 | 1.9 | 1.9 | 1.9 | 2.0 | 2.0 | 2.0 | 2.1 | 2.1 | 2.1 |
| 5 | 0.9 | 0.9 | 0.9 | 1.0 | 1.0 | 1.0 | 1.0 | 1.0 | 1.1 | 1.1 | 1.1 | 1.1 |
| 6 | 0.4 | 0.5 | 0.5 | 0.5 | 0.5 | 0.5 | 0.5 | 0.5 | 0.5 | 0.6 | 0.6 | 0.6 |
| 7 | 0.2 | 0.2 | 0.2 | 0.2 | 0.2 | 0.3 | 0.3 | 0.3 | 0.3 | 0.3 | 0.3 | 0.3 |
| 8+ | 0.2 | 0.2 | 0.2 | 0.2 | 0.2 | 0.2 | 0.2 | 0.2 | 0.2 | 0.3 | 0.3 | 0.3 |
| **Total (millions)** | **4.84** | **4.93** | **5.04** | **5.11** | **5.07** | **5.01** | **4.97** | **4.92** | **4.62** | **4.23** | **3.51** | **3.15** |

The proportion of the population with multimorbidity has increased by approximately 2.5 percentage points over the last decade, with more than 17% of all patients having at least two chronic morbidities in 2016. We find that the prevalence and the severity of multimorbidity increase progressively with age (see Figure 1). By age 60, approximately half the population has been diagnosed with at least one chronic condition, after which we observe a steep rise in multimorbidity, with close to 1 in 3 patients suffering from at least two morbidities by age 70. Stratifying the prevalence of multimorbidity by IMD in Figure 2, we find that people living in more deprived areas are more likely to be multimorbid compared to those living in more affluent areas at all ages. The same trend holds consistently for all years in our data.

**Figure 1.**
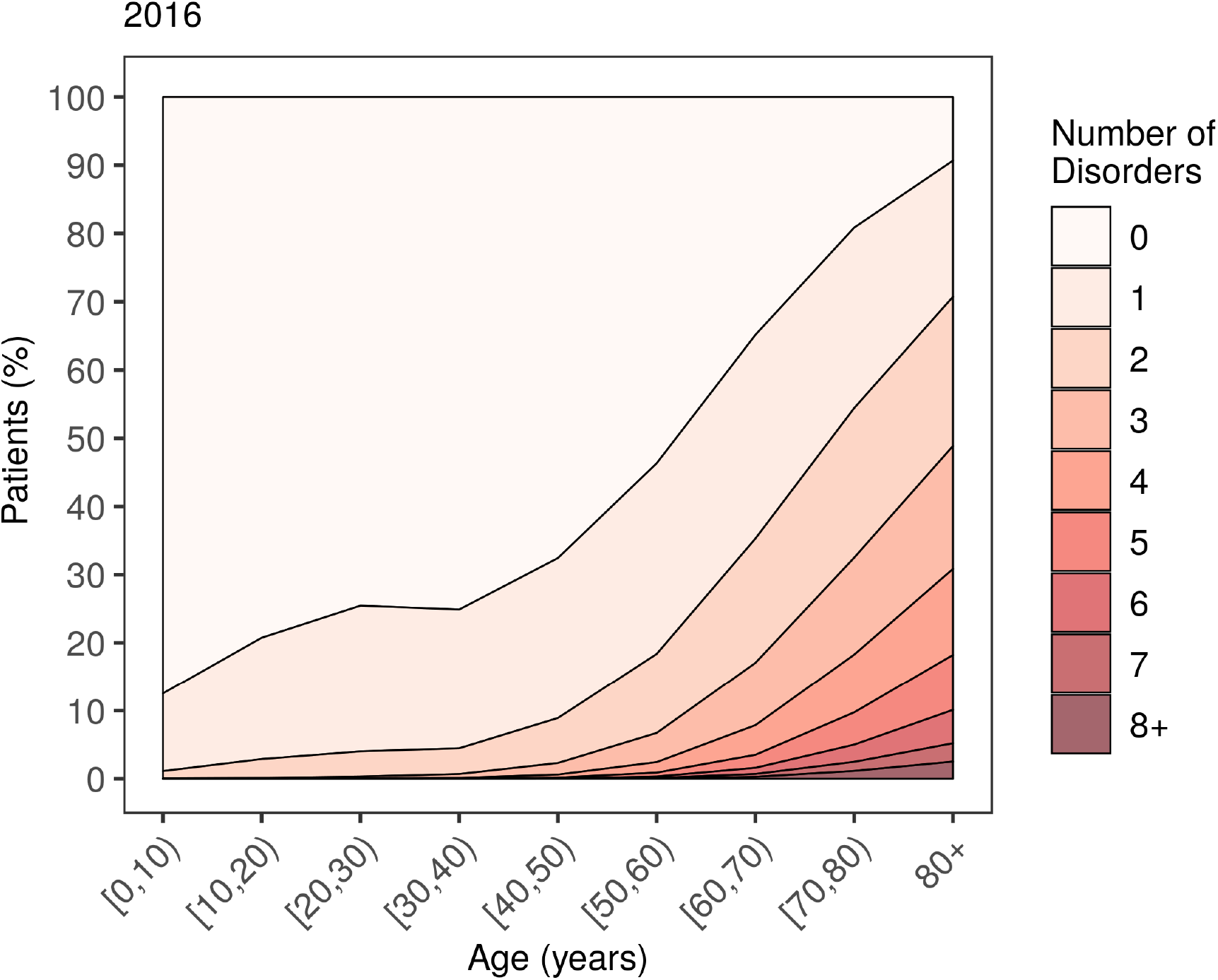
Number of co-occurring chronic conditions by age in 2016. See Appendix C for other years.

**Figure 2.**
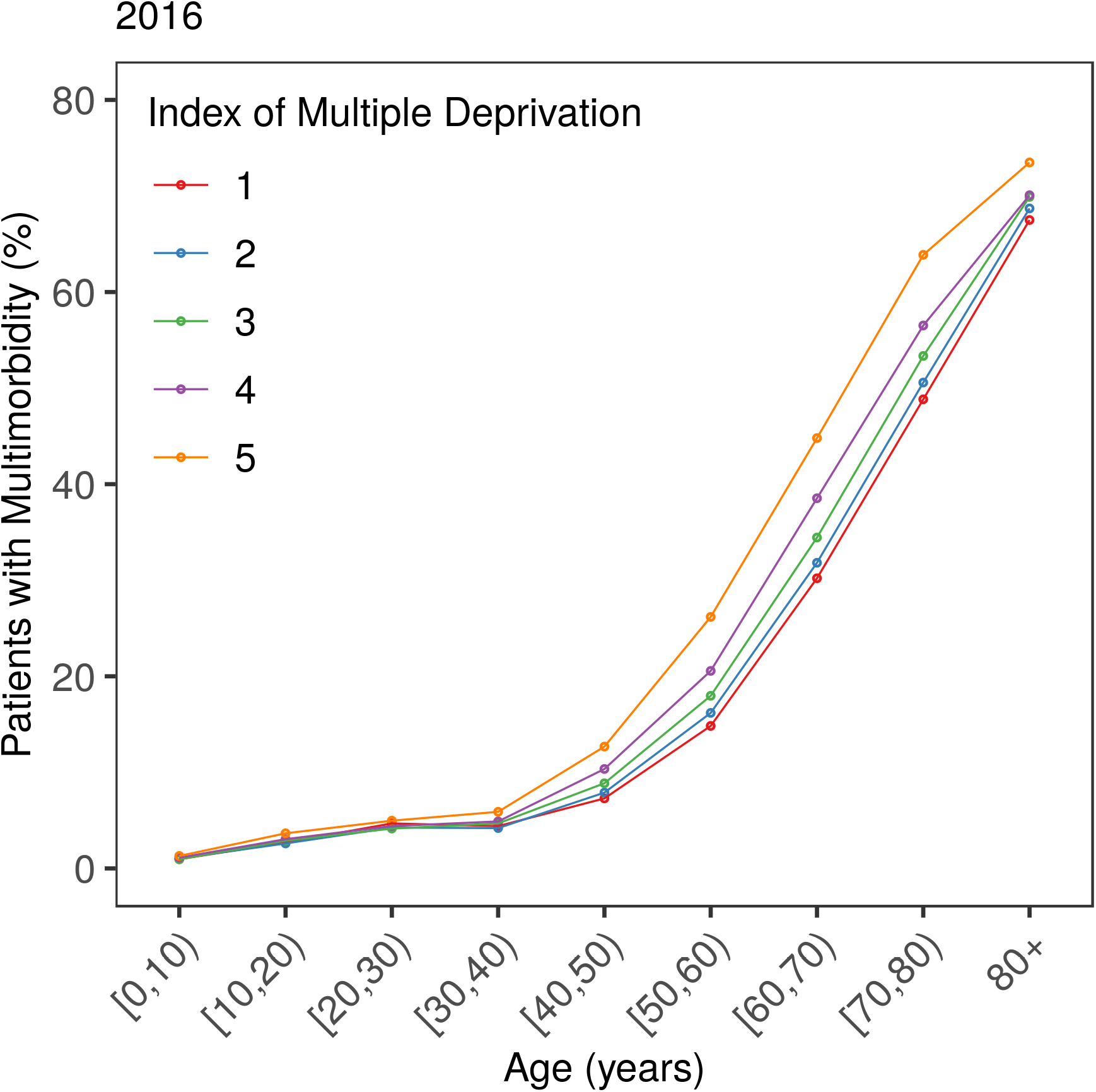
Prevalence of multimorbidity by age and IMD in 2016. See Appendix C for other years. Higher score corresponds to greater deprivation.

We characterize the epidemiology of individual diseases by plotting heat maps of disease prevalence in different age groups separately. We find that asthma and respiratory conditions have high prevalence across all age groups, with the former occurring especially frequently in the Adolescent age group (13 to 19 years old). We observe the onset of metabolic and cardiovascular diseases in the Middle-Aged and older age groups, in particular, of diabetes and hypertension. Not surprisingly, diseases such as dementia, kidney diseases, and stroke occur most frequently in the oldest patients (65 years and above). We observe an increasing trend in prevalence for some diseases. For example, the prevalence of diabetes in the Aged age group (65 to 80 years old) has increased by almost 35% in the past decade (see Figure 3). In contrast, the prevalence of diseases such as angina has been falling over the study period.

**Figure 3.**
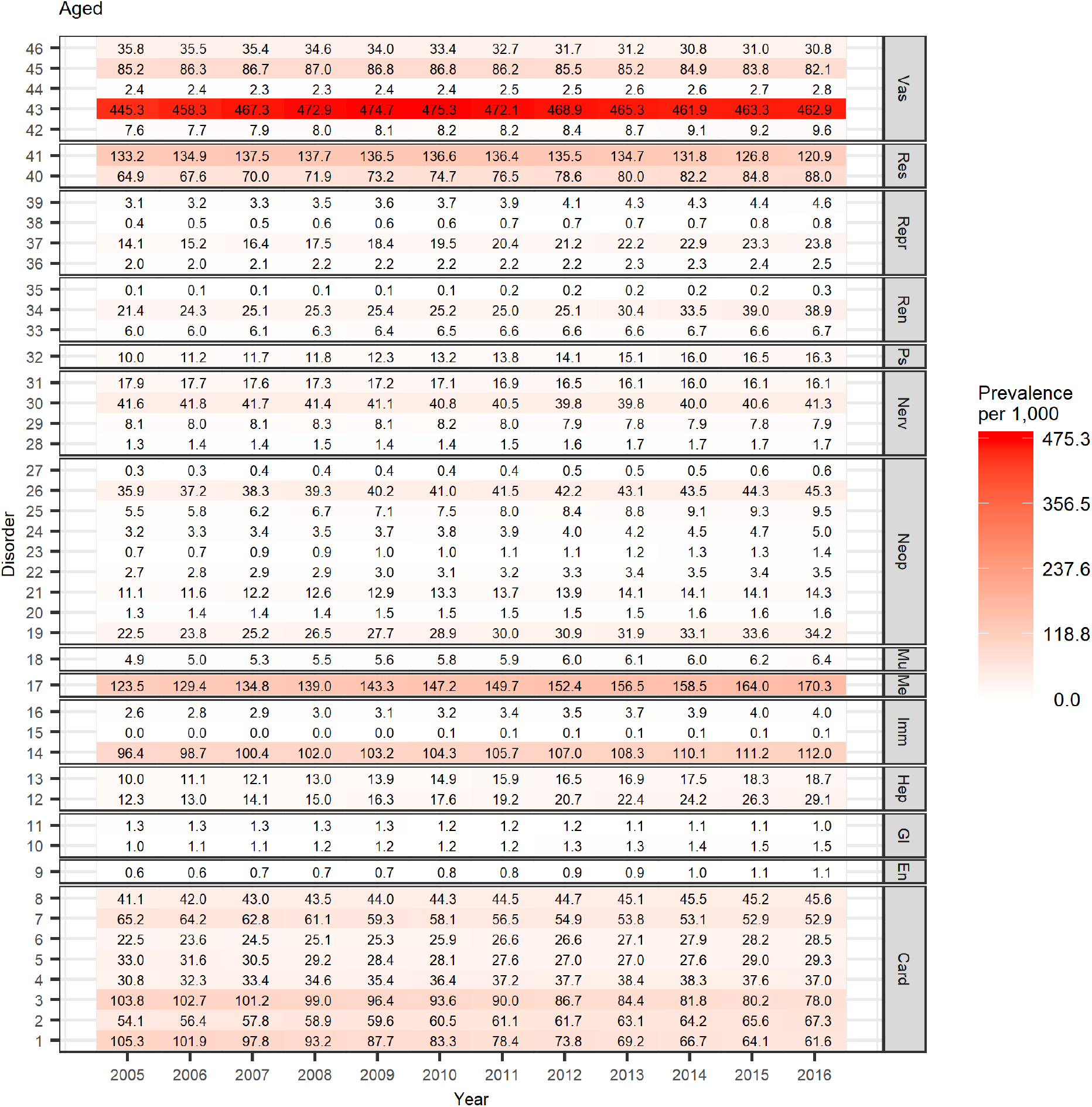
Single disease prevalence in the Aged subgroup between 2005 and 2016. See Figures 7 and 8 for disorder to index mapping and MedDRA SOC abbreviations. See Appendix C for other age groups.

In Table 2, we summarize the lift and odds ratio of the top ten most frequently co-occurring pairs of diseases in the Aged age group in 2016. (See Appendix C for other age groups and years.) In all age groups, asthma occurs in combination with other respiratory-related diseases approximately twice more often than expected by chance (i.e., the lift is greater than 2.0). Additionally, the estimated odds ratios, both unadjusted and adjusted, indicate that patients with asthma are at least twice as likely to suffer from other respiratory conditions at the same time, and vice versa.

**Table 2.** Lift and odds ratio of the top 10 most prevalent multimorbidity pairs in the Aged subgroup in 2016. See Appendix C for other age groups and years.

| Disease 1 | Disease 2 | N | Lift | Unadjusted OR (95% CI) | Adjusted OR (95% CI) |
| --- | --- | --- | --- | --- | --- |
| Diabetes | Hypertension | 50,105 | 1.5 | 3.0 (2.9, 3.0) | 2.7 (2.7, 2.8) |
| Hypertension | Other Respiratory Disease | 26,587 | 1.1 | 1.2 (1.2, 1.3) | 1.1 (1.1, 1.1) |
| Asthma | Hypertension | 24,320 | 1.1 | 1.2 (1.2, 1.2) | 1.1 (1.1, 1.2) |
| CAD | Hypertension | 19,479 | 1.2 | 1.6 (1.6, 1.7) | 1.2 (1.2, 1.2) |
| Hypertension | PAD | 19,203 | 1.2 | 1.4 (1.4, 1.4) | 1.2 (1.1, 1.2) |
| COPD | Hypertension | 18,529 | 1.1 | 1.1 (1.1, 1.1) | 0.9 (0.9, 1.0) |
| Atrial Fibrillation | Hypertension | 17,644 | 1.3 | 1.9 (1.8, 1.9) | 1.4 (1.4, 1.5) |
| Angina | Hypertension | 16,234 | 1.3 | 1.9 (1.8, 1.9) | 1.4 (1.4, 1.4) |
| Angina | CAD | 14,867 | 7.2 | 25.9 (25.2, 26.6) | 16.7 (16.2, 17.2) |
| Asthma | Other Respiratory Disease | 12,245 | 2.1 | 2.9 (2.8, 3.0) | 2.4 (2.4, 2.5) |

Hypertension is most associated with a second condition in the older age groups, although most pairs do not necessarily occur more frequently than by chance. The combination of hypertension and diabetes stands out with a relatively high lift and an odds ratio that is greater than 2.0, suggesting that this disease pair may be comorbid. Angina and coronary artery disease (CAD) also demonstrate a strong association in the Aged and Elderly age groups with unusually high lift and odds ratio.

To better visualize the data, we plot the lift of all combinations of disease pairs in heat maps, stratified by MedDRA system organ classes. (See Figure 4 and Appendix C for other age groups and years.) The co-occurrence of cardiac-cardiac and cardiac-respiratory disorders is a major risk across all age groups. We observe significant coupling between cardiac and hepatobiliary disorders in the Adolescent and Child (2 to 13 years old) age groups. On the other hand, combinations of cardiac-vascular and cardiac-metabolic disorders are the most dominant in the Middle-Aged and older age groups. We observe the same general patterns across time.

**Figure 4.**
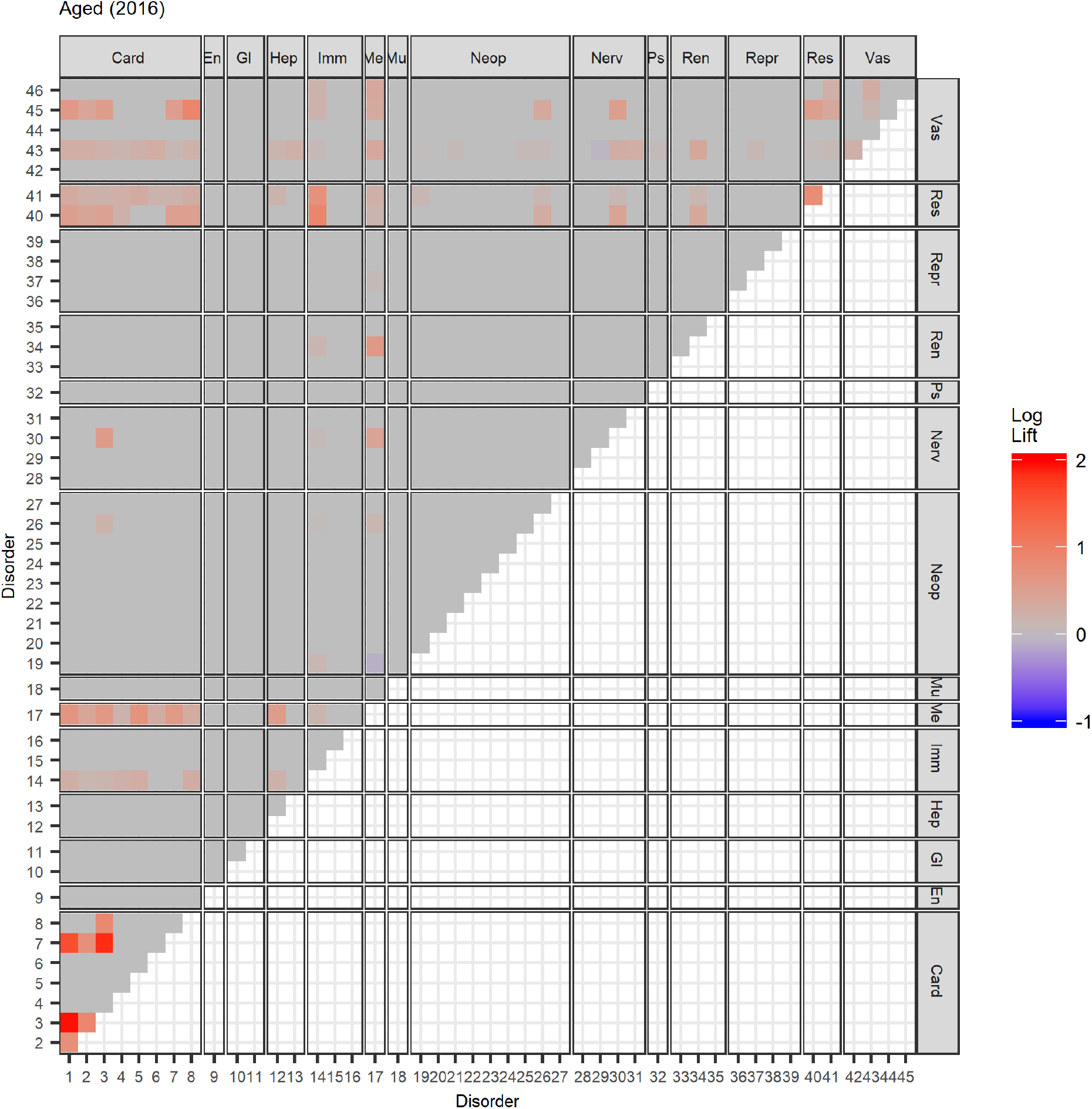
Heat map of lift of multimorbidity pairs in the Aged subgroup in 2016. See Figures 7 and 8 for disorder to index mapping and MedDRA SOC abbreviations. See Appendix C for other age groups and years.

As shown in Figure 1, the proportion of patients with three or more co-occurring disorders is small in the younger age groups. For patients aged 45 years and older, triplets involving angina, CAD, hypertension, diabetes and myocardial infarction (MI) occur most frequently with high lift, suggesting strong correlations between these diseases (see Table 3).

**Table 3.** Lift of the top 10 most prevalent multimorbidity triplets in the Aged subgroup in 2016. See Appendix C for other age groups and years.

| Disease 1 | Disease 2 | Disease 3 | N | Lift |
| --- | --- | --- | --- | --- |
| Angina | CAD | Hypertension | 8,988 | 9.3 |
| Diabetes | Hypertension | Other Respiratory Disease | 7,738 | 1.9 |
| CAD | Diabetes | Hypertension | 7,417 | 2.8 |
| Asthma | Diabetes | Hypertension | 6,761 | 1.8 |
| Asthma | Hypertension | Other Respiratory Disease | 6,457 | 2.4 |
| Angina | Diabetes | Hypertension | 6,304 | 3.0 |
| CAD | Hypertension | MI | 6,171 | 7.5 |
| Diabetes | Hypertension | PAD | 5,975 | 2.1 |
| Atrial Fibrillation | Diabetes | Hypertension | 5,417 | 2.4 |
| Asthma | COPD | Hypertension | 5,290 | 2.7 |

In Figures 5 and 6, we plot the undirected and directed multimorbidity networks observed in the Aged age group in 2016. (See Appendix C for other age groups and years.) Instead of a force-directed layout, we place the nodes in fixed positions around a circle to allow easy visualization of temporal changes in connections and clusters when comparing plots from different years. The edge thickness is proportional to the lift between each disease pair. Apart from single-node clusters, the communities detected using modularity maximization are given different colors.

**Figure 5.**
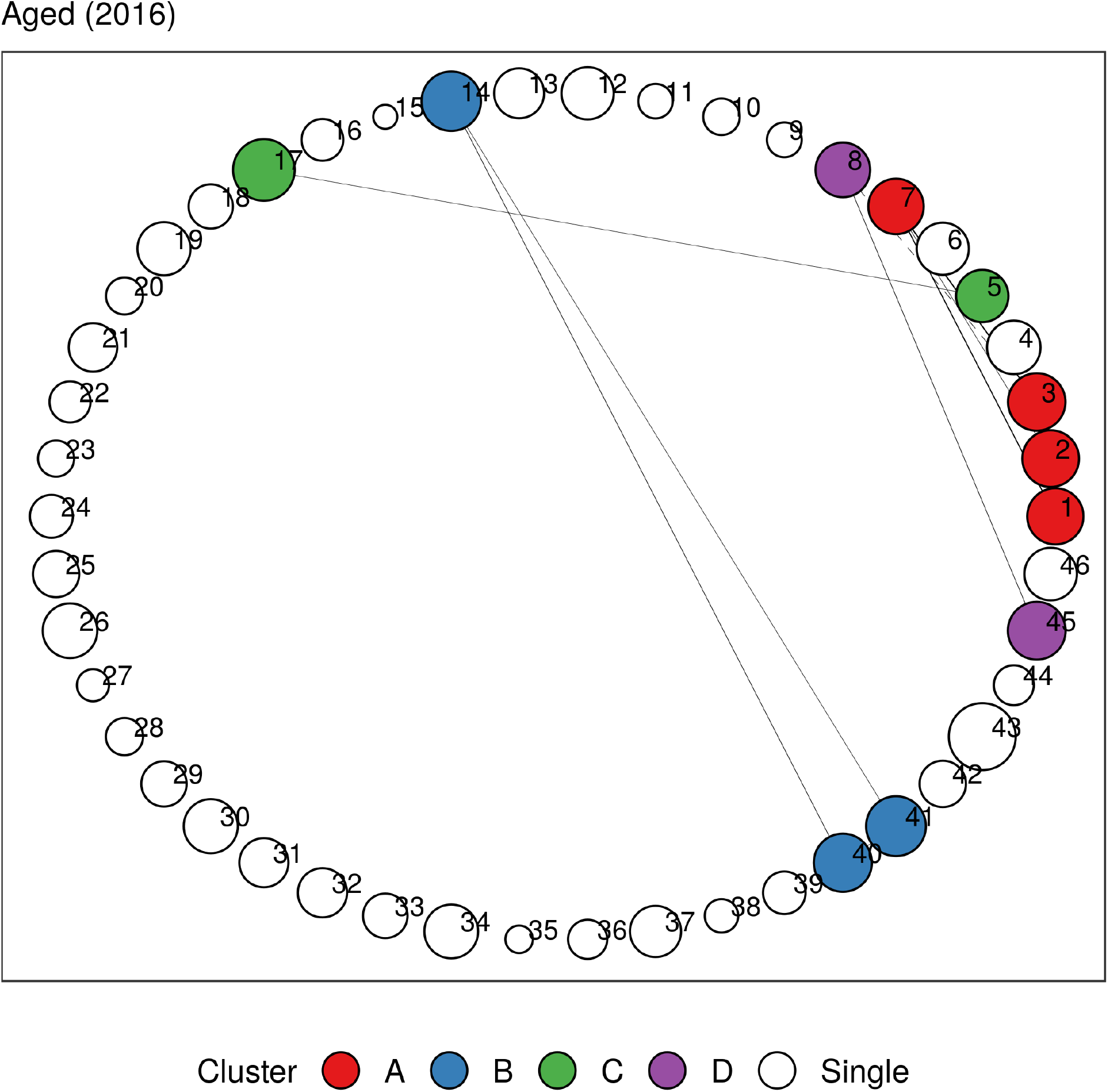
Undirected multimorbidity network in the Aged subgroup in 2016. Edge thickness is proportional to the lift between each disease pair. Intra-group edges and inter-group edges are represented by solid lines and dashed lines, respectively. Only communities with more than one node are colored. See Figures 7 and 8 for mapping of disorder to index and MedDRA SOC abbreviations. See Appendix C for other age groups and years.

**Figure 6.**
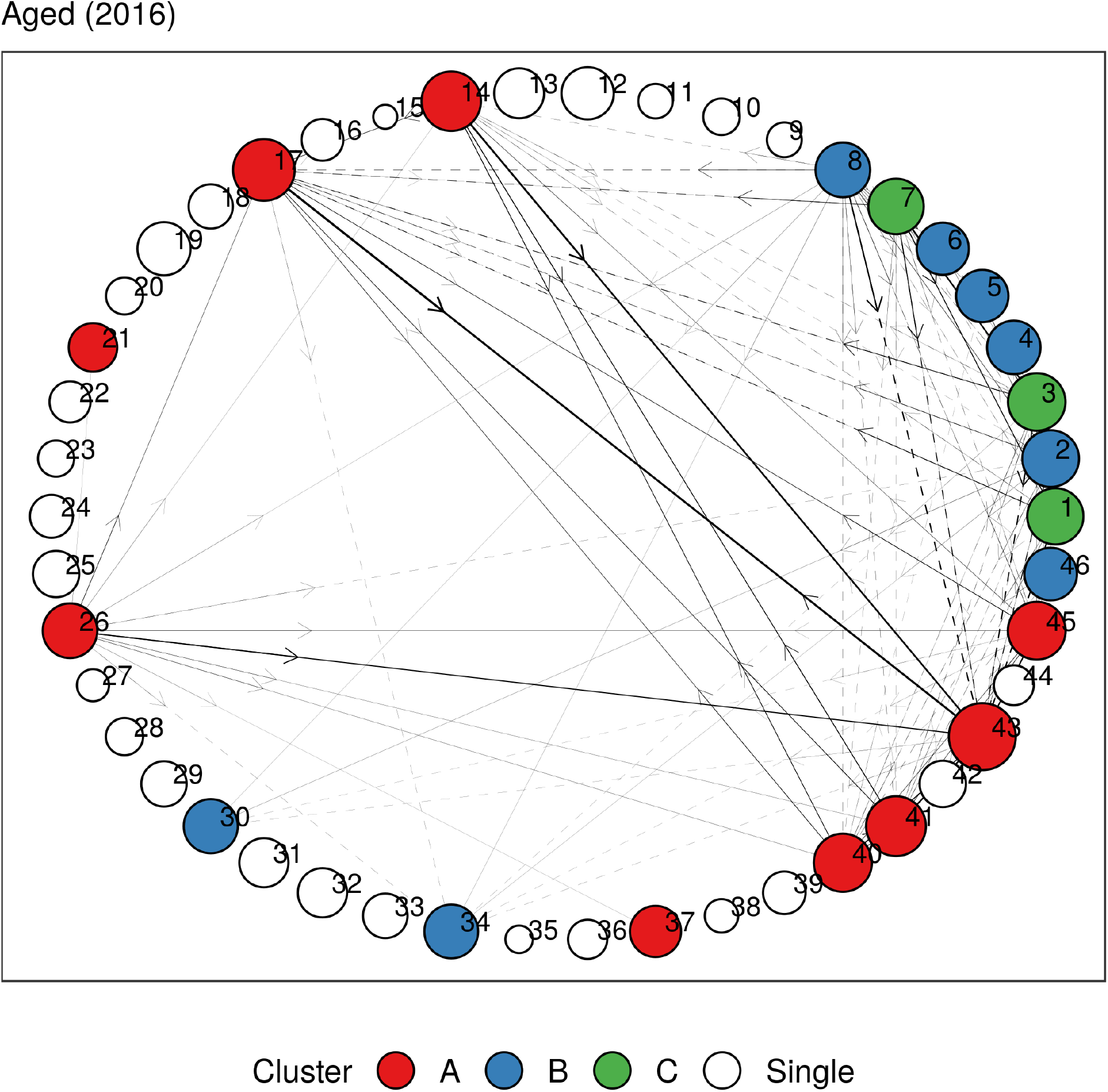
Directed multimorbidity network in the Aged subgroup in 2016. Edge thickness is proportional to the lift between each disease pair. Intra-group edges and inter-group edges are represented by solid lines and dashed lines, respectively. Only communities with more than one node are colored. See Figures 7 and 8 for mapping of disorder to index and MedDRA SOC abbreviations. See Appendix C for other age groups and years.

**Figure 7.**
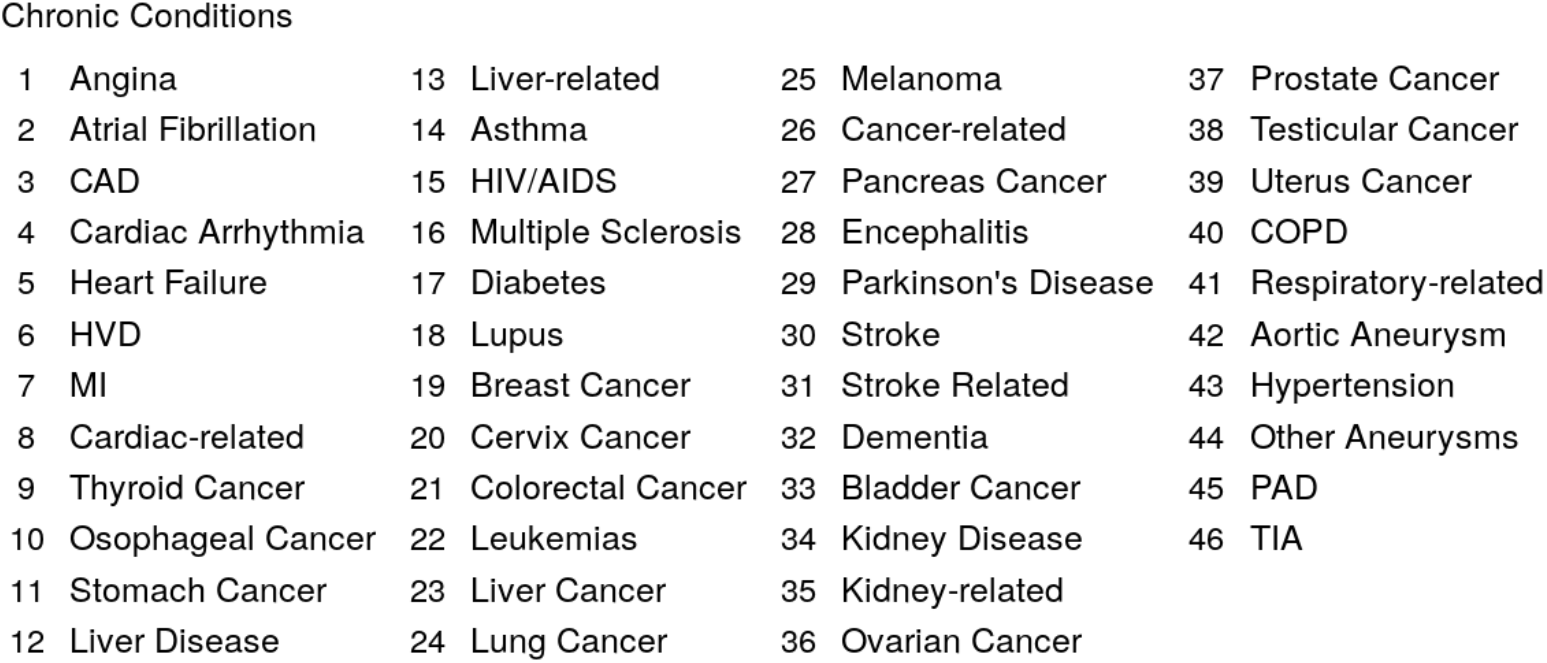
Mapping between index and chronic conditions.

**Figure 8.**
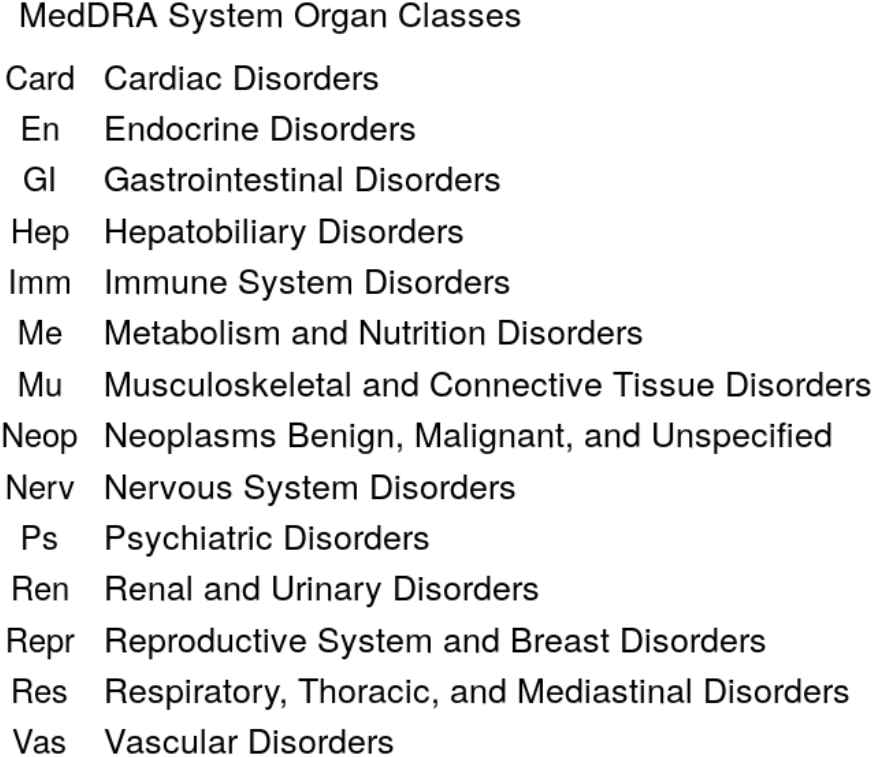
Abbreviations for MedDRA SOCs used in figures.

In Tables 4 and 5, we identify clusters that remain relatively stable throughout the years in undirected and directed multimorbidity networks, respectively. We find between one and four clusters for each age group. The number of diseases in each cluster ranges between two and twelve. In general, the communities found in Adolescent and younger patients can vary greatly from year to year compared to older age groups, where the clusters evolve very little over time. This is expected, given that only a small proportion of the former cohort suffers from more than two co-occurring disorders (see Figure 1), so the results are sensitive to small changes in prevalence each year.

**Table 4.** Clusters identified in undirected multimorbidity networks in different age groups.

| Age Group | Cluster 1 | Cluster 2 | Cluster 3 | Cluster 4 |
| --- | --- | --- | --- | --- |
| Infant | Asthma, COPD, Respiratory-related diseases | Cardiac arrhythmia, Heart failure, HVD, Cardiac-related diseases, Kidney disease, Hypertension |  |  |
| Child | Cardiac arrhythmia, HVD, Cardiac-related diseases, Hypertension | Liver disease, Liver-related diseases, Encephalitis, Stroke, Kidney disease, Hypertension, PAD |  |  |
| Adolescent | Asthma, COPD, Respiratory-related diseases | Cardiac arrhythmia, HVD, Cardiac-related diseases | Liver disease, Liver-related diseases, Diabetes, Leukemias, Kidney disease, Hypertension, PAD |  |
| Adult | Asthma, COPD, Respiratory-related diseases | Cardiac arrhythmia, Cardiac-related diseases, Kidney disease, PAD | HVD, Liver disease, Liver-related diseases, Diabetes, Lupus, Hypertension |  |
| Middle-Aged | CAD, MI, Cardiac-related diseases, Asthma, COPD, Respiratory-related diseases, PAD | Angina, Atrial fibrillation, Heart failure, Liver disease, Liver-related diseases, Diabetes, Stroke, Stroke-related diseases, Kidney disease, Hypertension, TIA |  |  |
| Aged | Cardiac-related diseases, PAD | Heart failure, Diabetes | Asthma, COPD, Respiratory-related diseases | Angina, Atrial fibrillation, CAD, Cardiac arrhythmia, MI, TIA |
| Elderly | Asthma, COPD, Respiratory-related diseases | Angina, CAD, MI | Atrial fibrillation, Cardiac arrhythmia, Heart failure, HVD |  |

**Table 5.** Clusters identified in directed multimorbidity networks in different age groups.

| Age Group | Cluster 1 | Cluster 2 | Cluster 3 | Cluster 4 |
| --- | --- | --- | --- | --- |
| Infant | Cardiac arrhythmia, Cardiac-related diseases, Liver disease, Liver-related diseases, Respiratory-related diseases |  |  |  |
| Child | Kidney disease, Hypertension, PAD | Cardiac arrhythmia, Heart failure, HVD, Cardiac-related diseases | Liver-related diseases, Asthma, Diabetes, Respiratory-related diseases |  |
| Adolescent | Liver-related diseases, Asthma, Respiratory-related diseases | Cardiac arrhythmia, Heart failure, HVD, Cardiac-related diseases | Diabetes, Kidney disease, Hypertension, PAD |  |
| Adult | Asthma, Respiratory-related diseases | Liver disease, Liver-related diseases | Diabetes, Lupus, Kidney disease, Kidney-related diseases, Hypertension | Atrial fibrillation, Heart failure, HVD, Cardiac-related diseases, Lupus, Stroke, PAD |
| Middle-Aged | Atrial fibrillation, Cardiac arrhythmia, HVD, Cardiac-related diseases, Stroke | Angina, CAD, Heart failure, MI, Stroke, TIA | Liver disease, Liver-related diseases, Asthma, Diabetes, Breast cancer, Colorectal cancer, Cancer-related diseases, Kidney disease, COPD, Respiratory-related diseases, Hypertension, PAD |  |
| Aged | Angina, CAD, MI | Atrial fibrillation, Cardiac arrhythmia, Heart failure, HVD, Cardiac-related diseases, Stroke, TIA | Asthma, Diabetes, Colorectal cancer, Cancer-related diseases, COPD, Respiratory-related diseases, Hypertension, PAD |  |
| Elderly | Angina, CAD, MI, Cardiac-related diseases | Asthma, Diabetes, COPD, Respiratory-related diseases, Hypertension, PAD | Atrial fibrillation, Cardiac arrhythmia, Heart failure, HVD, Stroke, Stroke-related diseases, Dementia, TIA |  |

A respiratory cluster of asthma, chronic obstructive pulmonary disease (COPD), and respiratory-related diseases appears to be present in all age groups in both undirected and directed graphs. Similarly, a vascular-metabolic-hepatobiliary-renal cluster that is characterized by hypertension, diabetes, liver diseases, and kidney diseases, with the occasional appearance of cardiac disorders, is also present in almost all cohorts. As observed in previous analyses, we also find several clusters dominated by cardiovascular disorders such as angina, CAD, myocardial infarction (MI), atrial fibrillation, cardiac arrhythmia, heart failure, heart valve disorder (HVD), stroke, peripheral artery disease (PAD), and transient ischemic attack (TIA).

In Tables 6 and 7, we summarize the top five diseases for each centrality measure. (See Appendix C for the full set of results.) The degree centrality and eigencentrality for hypertension, diabetes, CAD, and angina are the highest when all age groups are aggregated in undirected multimorbidity networks. In the Adolescent and younger age groups, kidney disease shows both high degree centrality and eigencentrality. Other important nodes include respiratory-related diseases and HVD, which have high degree centrality and high eigencentrality, respectively. For the Adult and Middle-Aged age groups, hypertension and diabetes are the most central nodes with respect to both measures. In the Aged and Elderly age groups, we find that cardiac disorders make up all of the top five most connected nodes.

**Table 6.** Centrality measures for top five diseases in undirected multimorbidity networks with mean computed over time.

| <u>All</u><br>Disease | Mean | <u>Infant</u><br>Disease | Mean | <u>Child</u><br>Disease | Mean | <u>Adolescent</u><br>Disease | Mean |
| --- | --- | --- | --- | --- | --- | --- | --- |
| <u>Degree Centrality</u> |  |  |  |  |  |  |  |
| Hypertension | 23.5 | Respiratory-related | 7.7 | Kidney Disease | 11.7 | Kidney Disease | 10.1 |
| Diabetes | 17.5 | Cardiac-related | 5.1 | Liver-related | 10.1 | Diabetes | 7.4 |
| PAD | 11.9 | HVD | 4.8 | Respiratory-related | 9.8 | Respiratory-related | 7.0 |
| CAD | 7.8 | Liver-related | 4.4 | HVD | 7.9 | Liver-related | 6.8 |
| Angina | 7.7 | Cardiac Arrhythmia | 3.7 | Cardiac-related | 6.5 | Asthma | 6.8 |
| <u>Eigencentality</u> |  |  |  |  |  |  |  |
| CAD | 0.9 | Cardiac-related | 0.9 | HVD | 1.0 | Kidney Disease | 1.0 |
| Diabetes | 0.9 | HVD | 0.8 | Kidney Disease | 0.9 | HVD | 0.9 |
| Hypertension | 0.9 | Hypertension | 0.6 | Cardiac-related | 0.8 | Cardiac-related | 0.9 |
| Angina | 0.9 | Cardiac Arrhythmia | 0.6 | Cardiac Arrhythmia | 0.8 | Liver Disease | 0.8 |
| PAD | 0.8 | Kidney Disease | 0.5 | Hypertension | 0.8 | Cardiac Arrhythmia | 0.7 |
| <u>Transitivity</u> |  |  |  |  |  |  |  |
|  | 0.32 |  | 0.29 |  | 0.37 |  | 0.40 |

**Table 6 (continued). Centrality measures for top five diseases in undirected multimorbidity networks with mean computed over time.**
| <u>Adult</u><br>Disease | Mean | <u>Middle-Aged</u><br>Disease | Mean | <u>Aged</u><br>Disease | Mean | <u>Elderly</u><br>Disease | Mean |
| --- | --- | --- | --- | --- | --- | --- | --- |
| <u>Degree Centrality</u> |  |  |  |  |  |  |  |
| Hypertension | 12.7 | Diabetes | 10.8 | CAD | 5.4 | Heart Failure | 3.5 |
| PAD | 8.5 | Hypertension | 9.5 | Angina | 4.7 | CAD | 3.1 |
| Cardiac-related | 7.8 | PAD | 6.8 | Atrial Fibrillation | 3.0 | Angina | 3.0 |
| Kidney Disease | 7.6 | COPD | 5.9 | MI | 3.0 | Atrial Fibrillation | 2.5 |
| Liver Disease | 6.7 | CAD | 5.7 | Cardiac-related | 2.8 | MI | 2.3 |
| <u>Eigencentality</u> |  |  |  |  |  |  |  |
| Hypertension | 1.0 | Diabetes | 0.8 | CAD | 1.0 | CAD | 1.0 |
| Cardiac-related | 0.8 | CAD | 0.8 | Angina | 0.9 | Angina | 0.9 |
| Liver Disease | 0.8 | Angina | 0.7 | MI | 0.8 | MI | 0.8 |
| Kidney Disease | 0.8 | Hypertension | 0.6 | Atrial Fibrillation | 0.5 | Heart Failure | 0.8 |
| Liver-related | 0.8 | PAD | 0.6 | Cardiac-related | 0.4 | Atrial Fibrillation | 0.4 |
| <u>Transitivity</u> |  |  |  |  |  |  |  |
|  | 0.66 |  | 0.31 |  | 0.59 |  | 0.57 |

**Table 7.** Centrality measures for top five diseases in directed multimorbidity networks with mean computed over time. We exclude eigencentralities that are close to zero.

| <u>All</u><br>Disease | Mean | <u>Infant</u><br>Disease | Mean | <u>Child</u><br>Disease | Mean | <u>Adolescent</u><br>Disease | Mean |
| --- | --- | --- | --- | --- | --- | --- | --- |
| <u>In-degree Centrality</u> |  |  |  |  |  |  |  |
| Diabetes | 11.0 | Respiratory-related | 5.3 | Asthma | 10.9 | Asthma | 11.1 |
| Respiratory-related | 11.0 | Cardiac-related | 0.5 | Respiratory-related | 10.7 | Respiratory-related | 10.9 |
| Hypertension | 11.0 | Kidney Disease | 0.4 | Kidney Disease | 3.4 | Hypertension | 3.9 |
| COPD | 10.8 | HVD | 0.3 | HVD | 3.1 | Kidney Disease | 3.1 |
| PAD | 10.1 | Liver-related | 0.3 | Cardiac Arrhythmia | 2.9 | HVD | 2.7 |
| <u>Out-degree Centrality</u> |  |  |  |  |  |  |  |
| CAD | 15.3 | Heart Failure | 1.3 | Cardiac-related | 6.3 | Hypertension | 7.5 |
| Atrial Fibrillation | 15.2 | Hypertension | 1.3 | Hypertension | 5.7 | Liver Disease | 6.5 |
| Angina | 14.9 | Liver-related | 1.0 | Liver Disease | 5.3 | HVD | 6.0 |
| Cardiac-related | 14.0 | Stroke | 0.9 | Leukemias | 5.2 | Cardiac-related | 5.8 |
| Hypertension | 12.9 | Cardiac Arrhythmia | 0.7 | HVD | 4.4 | Cancer-related | 3.8 |
| <u>Eigencentrality</u> |  |  |  |  |  |  |  |
| Hypertension | 1.0 |  |  | Asthma | 0.9 | Asthma | 0.9 |
| Diabetes | 0.4 |  |  | Respiratory-related | 0.4 | Respiratory-related | 0.4 |
| CAD | 0.4 |  |  |  |  |  |  |
| Respiratory-related | 0.4 |  |  |  |  |  |  |
| Angina | 0.3 |  |  |  |  |  |  |
| <u>Transitivity</u> |  |  |  |  |  |  |  |
|  | 0.76 |  | 0.14 |  | 0.57 |  | 0.57 |

**Table 7 (continued). Centrality measures for top five diseases in directed multimorbidity networks with mean computed over time. We exclude eigencentralities that are close to zero.**
| <u>Adult</u> |  | <u>Middle-Aged</u> |  | <u>Aged</u> |  | <u>Elderly</u> |  |
| --- | --- | --- | --- | --- | --- | --- | --- |
| Disease | Mean | Disease | Mean | Disease | Mean | Disease | Mean |
| <u>In-degree Centrality</u> |  |  |  |  |  |  |  |
| Asthma | 11.0 | Asthma | 11.0 | Atrial Fibrillation | 11.0 | Atrial Fibrillation | 11.0 |
| Diabetes | 11.0 | Diabetes | 11.0 | Diabetes | 11.0 | Hypertension | 11.0 |
| Respiratory-related | 11.0 | Respiratory-related | 11.0 | Respiratory-related | 11.0 | Diabetes | 10.6 |
| Hypertension | 11.0 | Hypertension | 11.0 | Hypertension | 11.0 | PAD | 10.4 |
| PAD | 11.0 | PAD | 11.0 | PAD | 11.0 | CAD | 10.3 |
| <u>Out-degree Centrality</u> |  |  |  |  |  |  |  |
| Cardiac-related | 13.4 | Cardiac-related | 16.8 | Cardiac-related | 14.2 | CAD | 13.1 |
| Kidney Disease | 12.3 | CAD | 14.6 | CAD | 13.8 | Hypertension | 12.5 |
| HVD | 12.3 | Hypertension | 14.5 | Atrial Fibrillation | 13.1 | Angina | 12.4 |
| Cancer-related | 10.4 | PAD | 10.9 | Angina | 12.0 | PAD | 12.4 |
| Hypertension | 10.3 | MI | 10.7 | Hypertension | 11.7 | Atrial Fibrillation | 12.3 |
| <u>Eigencentrality</u> |  |  |  |  |  |  |  |
| Asthma | 1.0 | Hypertension | 1.0 | Hypertension | 1.0 | Hypertension | 1.0 |
| Respiratory-related | 0.7 | Diabetes | 0.5 | Diabetes | 0.4 | CAD | 0.4 |
| Hypertension | 0.5 | Respiratory-related | 0.4 | CAD | 0.4 | Angina | 0.3 |
| Diabetes | 0.3 | Asthma | 0.3 | Angina | 0.3 | Atrial Fibrillation | 0.3 |
| PAD | 0.2 | CAD | 0.3 | Respiratory-related | 0.3 | Diabetes | 0.3 |
| <u>Transitivity</u> |  |  |  |  |  |  |  |
|  | 0.82 |  | 0.70 |  | 0.78 |  | 0.75 |

We observe similar results in directed networks. In general, hypertension, diabetes, and respiratory-related diseases demonstrate high in-degree centrality and eigencentrality, while cardiac disorders show high out-degree centrality. In the Middle-Aged and younger age groups, asthma emerges as a new central node with high in-degree centrality, while the top five diseases for the Aged and Elderly age groups remain dominated by cardiovascular diseases.

We summarize the dataset used for survival analysis in Table 8. The sample consists of approximately 390,000 patients in the Aged and Elderly age groups for each year between 2010 and 2012. More than 50% of the patients are multimorbid. In terms of predicting five-year overall survival, we find the performance of the linear and nonlinear survival models explored to be very similar (see Appendix D). We focus on the Cox model here due to its ease of interpretability. The model achieves a promising C-index of 0.81 (95% CI 0.80–0.81) on out-of-sample data in 2012. In addition, its calibration curves lay close to the ideal diagonal, indicating that the model is well calibrated, i.e., the model does not systematically overestimate or underestimate survival rates in any of the quintiles. (See Appendix C for plots.)

**Table 8.**
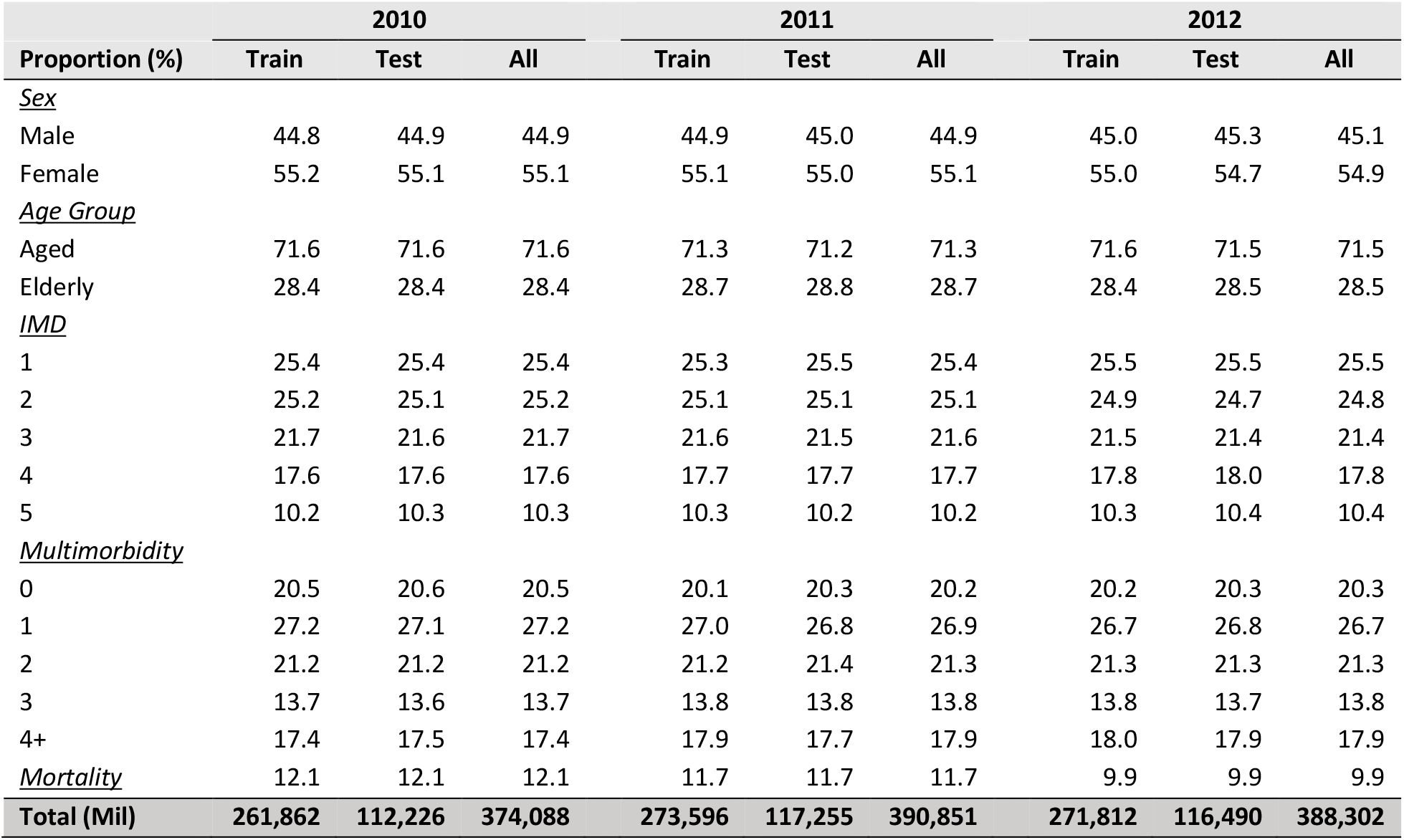
Summary statistics of dataset used for survival analysis.

In Table 9, we extract the top ten coefficients in the Cox model to identify specific risk factors. To correct for multiple testing, we perform the Benjamini-Hochberg adjustment with a 5% false discovery rate for identifying significant factors. Apart from cancers, we find the presence of multimorbidity to be a strong adverse risk factor, i.e., the higher the number of co-occurring chronic conditions, the greater the mortality risk. For example, the hazard ratio of having four or more chronic conditions is 2.44 (95% CI 2.22–2.69). We also find a high IMD, corresponding to a lower socioeconomic status, to be significantly associated with increased risk, although this factor is not in the top ten coefficients.

**Table 9.** Top ten risk factors of the Cox Proportional Hazards model for 2012. Asterisks denote factors that are significant after the Benjamini-Hochberg adjustment.

| Factor | HR (95% CI) |  |
| --- | --- | --- |
| Pancreas Cancer | 4.67 (3.61, 6.02) | * |
| Osophageal Cancer | 4.42 (3.75, 5.22) | * |
| Lung Cancer | 4.38 (3.99, 4.80) | * |
| Liver Cancer | 4.35 (3.66, 5.16) | * |
| Multimorbidity (4+) | 2.44 (2.22, 2.69) | * |
| Dementia | 2.28 (2.18, 2.39) | * |
| Ovarian Cancer | 2.21 (1.84, 2.65) | * |
| Multimorbidity (3) | 2.16 (2.00, 2.34) | * |
| Parkinson's Disease | 2.06 (1.91, 2.23) | * |
| Multimorbidity (2) | 1.89 (1.76, 2.02) | * |

## 4 Discussion

With multimorbidity becoming the norm rather than the exception^2,12,17,22,42,43^, the management of multiple chronic diseases in older adults is a major challenge facing healthcare systems worldwide. It is clear that a better understanding of the epidemiology of multimorbidity is required to develop more effective preventive interventions and better primary medical care for multimorbid patients. In this paper, we use data-driven methods to characterize multimorbidity patterns in different demographic groups and their evolution over the past decade, using a large, representative electronic medical records database consisting of over 4.5 million patients.

Consistent with other studies, we find that the prevalence and severity of multimorbidity increase substantially with age. In addition, we observe social inequalities in multimorbidity, with patients in socioeconomically deprived areas more likely to be multimorbid.^11,12,31,42,44– 46^ Our findings also support the role of hypertension as an important risk factor in older adults, as reported in the literature.^2,33,47^ Hypertension is one of the most prevalent and most central chronic conditions in our dataset, and one that serves as an important bridge between many diseases in our networks. Other trends identified in our analysis, such as the falling prevalence of angina^48–51^ and the growing prevalence of diabetes^52^, are also well documented in previously published population studies.

In our pairwise analysis, we find strong association between multiple pairs of chronic conditions, including between asthma and respiratory-related diseases^53,54^ in the Adolescent age group, between hypertension and diabetes^25,55–57^ and between CAD and angina^58^ among older patients, and between cardiovascular and respiratory disorders in all age groups.^2^ Triplets involving cardiovascular and metabolic disorders, such as CAD, hypertension, and diabetes, also occurred more frequently than expected by chance.^2,22,25,59^

Our network analysis further identified several meaningful communities that are common across all demographics, including a respiratory cluster (e.g., asthma and COPD)^60^, a cardiovascular cluster^19,61^, and a mixed cardiovascular-renal-metabolic cluster^32,62–64^, all of which are supported by either established pathophysiological mechanisms or shared risk factors. For example, it is well known that cardiovascular diseases are one of the most common complications of diabetes. While we do not find any particular multimorbidity pattern to have a significant effect on mortality, our models do indeed verify the substantial burden of multimorbidity (as quantified by the number of co-occurring chronic conditions) on overall survival in older patients.^7,12,23,65,66^

However, we must emphasize that our results do not necessarily imply any causal link between diseases identified to be in the same cluster. The association might be attributable to shared risk factors (e.g., smoking) or other adverse events, and any temporal relationships to be inferred from the multimorbidity directed networks might be administrative in nature (e.g., incomplete medical records that are rectified in subsequent visits) or biased by delayed diagnosis.

In general, the lack of an accepted standard for defining multimorbidity makes it difficult for any meaningful comparison of results across different studies.^67,687^ Moreover, because results can be highly dependent on the study population, the disease ontology used, and the number of chronic conditions considered, it is not uncommon for studies to report seemingly conflicting findings. In this paper, we consider a wide range of demographic groups and a total of 46 morbidities, which is more than most similar studies,^11^ and well above the minimum of 11 to 12 as recommended by systematic reviews in this field of research.^68,69^ In addition, our findings are largely consistent with existing studies in the medical literature.

Lastly, we note that cancer appears to be under represented in the THIN database. This is because many cancer patients are treated separately in cancer centers under the care of specialized clinical teams. Unfortunately, data on such patients rarely make their way back to the primary care clinics where the THIN data is collected, leading to a gap in this area.

## 5 Conclusions

Current healthcare systems are largely centered on single-disease approaches to treatment, resulting in the fragmentation of care and a lack of continuity in the management of multiple diseases. Even most clinical trials exclude multimorbid patients. Because multimorbidity is more common in disadvantaged groups, the current structure exacerbates health inequalities in society. In this paper, we apply statistical methods and network analysis to characterize multimorbidity associations in the general UK population using a large electronic medical records database spanning the years 2005 to 2016. Our findings contribute to a better understanding of multimorbidity that may be useful for the early detection and prevention of comorbidities, for example, recommending that hypertension patients reduce sugar intake as a preventive measure for diabetes.

There is a pressing need for a universal framework that standardizes the way that multimorbidity is assessed (e.g., the minimum number of diseases and the choice of chronic conditions to include) in order to facilitate comparisons between studies and populations. With the “Omics” revolution, the combination of phenotypic, genomic, and epigenomic data has the potential to provide deeper insights into the underlying pathophysiological associations between comorbid diseases. Unfortunately, the availability of such linked datasets remains very limited. Further research is also needed to better understand the impact of multimorbidity on different health outcomes, such as the quality of life and healthcare costs, in order to align the primary healthcare system more closely to the needs to multimorbid patients.

## Data Availability

The data supporting the current study have not been deposited in a public repository due to its proprietary nature. The data are available from The Health Improvement Network (THIN) at https://www.the-health-improvement-network.com/en/. Restrictions apply to the availability of the data, which were used under license for this study.

## Declarations

### Ethics approval and consent to participate

This study was approved by the Scientific Review Committee at IQVIA.

### Consent for publication

Not applicable

### Availability of data and materials

The data that support the findings of this study are available from The Health Improvement Network (THIN; https://www.the-health-improvement-network.com/en/) but restrictions apply to the availability of these data, which were used under license for the current study, and so are not publicly available. Data are however available from the authors upon reasonable request and with permission of THIN.

### Competing interests

K.W.S. and C.H.W. declare no competing interests. J.G. is an employee of Swiss Re and declares no competing interests. A.L. reports personal investments in private biotech companies, biotech venture capital funds, and mutual funds. A.L. is a co-founder and partner of QLS Advisors, a healthcare analytics and consulting company; an advisor to BrightEdge Ventures; a director of BridgeBio Pharma, Roivant Sciences, and Annual Reviews; chairman emeritus and senior advisor to AlphaSimplex Group; and a member of the Board of Overseers at Beth Israel Deaconess Medical Center and the NIH’s National Center for Advancing Translational Sciences Advisory Council and Cures Acceleration Network Review Board. During the most recent six-year period, A.L. has received speaking/consulting fees, honoraria, or other forms of compensation from: AIG, AlphaSimplex Group, BIS, BridgeBio Pharma, Citigroup, Chicago Mercantile Exchange, Financial Times, FONDS Professionell, Harvard University, IMF, National Bank of Belgium, Q Group, Roivant Sciences, Scotia Bank, State Street Bank, University of Chicago, and Yale University.

### Funding

No direct funding was received for this study; general research support was provided by the MIT Laboratory for Financial Engineering and its sponsors. The authors were personally salaried by their institutions during the period of writing (though no specific salary was set aside or given for the writing of this paper).

### Author contributions

Conceptualization, K.W.S., C.H.W., J.G., and A.L.; Resources, J.G., and A.L.; Methodology, K.W.S., C.H.W., J.G., and A.L.; Software, K.W.S.; Formal Analysis, K.W.S.; Writing – Original Draft, K.W.S. and A.L.; Writing – Review & Editing, K.W.S., C.H.W., J.G., and A.L.; Supervision, A.L.; Project Administration, J.G., and A.L.

## Acknowledgments

Research support from the MIT Laboratory for Financial Engineering is gratefully acknowledged. The views and opinions expressed in this article are those of the authors only, and do not necessarily represent the views and opinions of any institution or agency, any of their affiliates or employees, or any of the individuals acknowledged above.

## Notes

### Author Declarations

This study was approved by the Scientific Review Committee at IQVIA.

## References

1. World Health Organization. Multimorbidity Technical Series on Safer Primary Care Multimorbidity: Technical Series on Safer Primary Care. (2016).

2. Schäfer, I. et al.. Reducing complexity: A visualisation of multimorbidity by combining disease clusters and triads. BMC Public Health 14, 1285 (2014).

3. Kadam, U. & Croft, P. Clinical multimorbidity and physical function in older adults: a record and health status linkage study in general practice. Fam. Pract. 24, 412–419 (2007).

4. Laux, G., Kuehlein, T., Rosemann, T. & Szecsenyi, J. Co- and multimorbidity patterns in primary care based on episodes of care: Results from the German CONTENT project. BMC Health Serv. Res. 8, 14 (2008).

5. Fung, C. H. et al.. The relationship between multimorbidity and patients’ ratings of communication. J. Gen. Intern. Med. 23, 788–793 (2008).

6. Schoenberg, N. E., Kim, H., Edwards, W. & Fleming, S. T. Burden of Common Multiple-Morbidity Constellations on Out-of-Pocket Medical Expenditures Among Older Adults. Gerontologist 47, 423–437 (2007).

7. Gijsen, R. et al.. Causes and consequences of comorbidity: A review. J. Clin. Epidemiol. 54, 661–674 (2001).

8. Salisbury, C., Johnson, L., Purdy, S., Valderas, J. M. & Montgomery, A. A. Epidemiology and impact of multimorbidity in primary care: a retrospective cohort study. Br. J. Gen. Pract. 61, e12–e21 (2011).

9. Wolff, J. L., Starfield, B. & Anderson, G. Prevalence, Expenditures, and Complications of Multiple Chronic Conditions in the Elderly. Arch. Intern. Med. 162, 2269 (2002).

10. Fortin, M. et al.. Multimorbidity and quality of life in primary care: A systematic review. Health Qual. Life Outcomes 2, 1–12 (2004).

11. Barnett, K. et al.. Epidemiology of multimorbidity and implications for health care, research, and medical education: A cross-sectional study. Lancet 380, 37–43 (2012).

12. Marengoni, A. et al.. Aging with multimorbidity: A systematic review of the literature. Ageing Res. Rev. 10, 430–439 (2011).

13. Loza, E., Jover, J. A., Rodriguez, L. & Carmona, L. Multimorbidity: Prevalence, Effect on Quality of Life and Daily Functioning, and Variation of This Effect When one Condition Is a Rheumatic Disease. Semin. Arthritis Rheum. 38, 312–319 (2009).

14. Crentsil, V., Ricks, M. O., Xue, Q. L. & Fried, L. P. A pharmacoepidemiologic study of community-dwelling, disabled older women: Factors associated with medication use. Am. J. Geriatr. Pharmacother. 8, 215–224 (2010).

15. Walker, A. E. Multiple chronic diseases and quality of life: Patterns emerging from a large national sample, Australia. Chronic Illn. 3, 202–218 (2007).

16. Van den Akker, M., Buntix, F., Metsemakers, J. F. M., Roos, S. & Knottnerus, J. A. Multimorbidity in general practice: Prevalence, incidence, and determinants of co-occurring chronic and recurrent diseases. J. Clin. Epidemiol. 51, 367–375 (1998).

17. Fortin, M. Prevalence of Multimorbidity Among Adults Seen in Family Practice. Ann. Fam. Med. 3, 223–228 (2005).

18. Boyd, C. M. et al.. Clinical Practice Guidelines and Quality of Care for Older Patients With Multiple Comorbid Diseases. JAMA 294, 716 (2005).

19. Marengoni, A., Rizzuto, D., Wang, H.-X., Winblad, B. & Fratiglioni, L. Patterns of Chronic Multimorbidity in the Elderly Population. J. Am. Geriatr. Soc. 57, 225–230 (2009).

20. Marengoni, A. et al.. In-hospital death and adverse clinical events in elderly patients according to disease clustering: The REPOSI study. Rejuvenation Res. 13, 469–477 (2010).

21. Redelmeier, D. A., Tan, S. H. & Booth, G. L. The Treatment of Unrelated Disorders in Patients with Chronic Medical Diseases. N. Engl. J. Med. 338, 1516–1520 (1998).

22. Schäfer, I. et al.. Multimorbidity Patterns in the Elderly: A New Approach of Disease Clustering Identifies Complex Interrelations between Chronic Conditions. PLoS One 5, e15941 (2010).

23. Ferrer, A., Formiga, F., Sanz, H., Almeda, J. & Padrós, G. Multimorbidity as specific disease combinations, an important predictor factor for mortality in octogenarians: the Octabaix study. Clin. Interv. Aging 12, 223–231 (2017).

24. Diederichs, C., Berger, K. & Bartels, D. B. The Measurement of Multiple Chronic Diseases--A Systematic Review on Existing Multimorbidity Indices. Journals Gerontol. Ser. A Biol. Sci. Med. Sci. 66A, 301–311 (2011).

25. Kirchberger, I. et al.. Patterns of Multimorbidity in the Aged Population. Results from the KORA-Age Study. PLoS One 7, e30556 (2012).

26. The Health Improvement Network. The Health Improvement Network. https://www.the-health-improvement-network.com/en/.

27. Kastner, M., Wilczynski, N. L., Walker-Dilks, C., McKibbon, K. A. & Haynes, B. Agespecific search strategies for medline. J. Med. Internet Res. 8, (2006).

28. National Health Services. Read Codes. https://digital.nhs.uk/services/terminology-and-classifications/read-codes (2020).

29. Timmreck, T. C., Cole, G. E., James, G. & Butterworth, D. D. Health education and health promotion: A look at the jungle of supportive fields, philosophies and theoretical foundations. Health Educ. 18, 23–28 (1987).

30. Ministry of Housing Communities & Local Government. English indices of deprivation. https://www.gov.uk/government/collections/english-indices-of-deprivation (2012).

31. Orueta, J. F., García-Álvarez, A., García-Goñi, M., Paolucci, F. & Nuño-Solinís, R. Prevalence and Costs of Multimorbidity by Deprivation Levels in the Basque Country: A Population Based Study Using Health Administrative Databases. PLoS One 9, e89787 (2014).

32. Aguado, A., Moratalla-Navarro, F., López-Simarro, F. & Moreno, V. MorbiNet: multimorbidity networks in adult general population. Analysis of type 2 diabetes mellitus comorbidity. Sci. Rep. 10, 2416 (2020).

33. Feldman, K. et al.. Insights into Population Health Management Through Disease Diagnoses Networks. Sci. Rep. 6, 30465 (2016).

34. Leva, F. & Bitonti, D. Network analysis of comorbidity patterns in heart failure patients using administrative data. Epidemiol. Biostat. Public Heal. 15, (2018).

35. Liu, J. et al.. Comorbidity Analysis According to Sex and Age in Hypertension Patients in China. Int. J. Med. Sci. 13, 99–107 (2016).

36. Brandes, U. et al.. On modularity clustering. IEEE Trans. Knowl. Data Eng. 20, 172–188 (2008).

37. Girvan, M. & Newman, M. E. J. Community structure in social and biological networks. Proc. Natl. Acad. Sci. 99, 7821–7826 (2002).

38. Newman, M. E. J. Fast algorithm for detecting community structure in networks. Phys. Rev. E - Stat. Physics, Plasmas, Fluids, Relat. Interdiscip. Top. 69, 5 (2004).

39. Cox, D. R. Regression Models and Life-Tables. J. R. Stat. Soc. Ser. B 34, 187–202 (1972).

40. Faraggi, D. & Simon, R. A neural network model for survival data. Stat. Med. 14, 73–82 (1995).

41. Harrell, F. E. Evaluating the Yield of Medical Tests. JAMA J. Am. Med. Assoc. 247, 2543 (1982).

42. Violan, C. et al.. Prevalence, Determinants and Patterns of Multimorbidity in Primary Care: A Systematic Review of Observational Studies. PLoS One 9, e102149 (2014).

43. Boyd, C. M. & Fortin, M. Future of multimorbidity research: How should understanding of multimorbidity inform health system design? Public Health Rev. 32, 451–474 (2010).

44. Schiøtz, M. L., Stockmarr, A., Høst, D., Glümer, C. & Frølich, A. Social disparities in the prevalence of multimorbidity - A register-based population study. BMC Public Health 17, 422 (2017).

45. Dugravot, A. et al.. Social inequalities in multimorbidity, frailty, disability, and transitions to mortality: a 24-year follow-up of the Whitehall II cohort study. Lancet Public Heal. 5, e42–e50 (2020).

46. Schäfer, I. et al.. The influence of age, gender and socio-economic status on multimorbidity patterns in primary care. first results from the multicare cohort study. BMC Health Serv. Res. 12, 89 (2012).

47. Chen, Y. & Xu, R. Mining Cancer-Specific Disease Comorbidities from a Large Observational Health Database. Cancer Inform. 13s1, CIN.S13893 (2014).

48. Abdalla, S. M., Yu, S. & Galea, S. Trends in Cardiovascular Disease Prevalence by Income Level in the United States. JAMA Netw. Open 3, e2018150 (2020).

49. Yoon, S. S. (Sarah), Dillon, C. F., Illoh, K. & Carroll, M. Trends in the Prevalence of Coronary Heart Disease in the U.S.: National Health and Nutrition Examination Survey, 2001–2012. Am. J. Prev. Med. 51, 437–445 (2016).

50. Benjamin, E. J. et al.. Heart Disease and Stroke Statistics—2019 Update: A Report From the American Heart Association. Circulation 139, e56–e528 (2019).

51. Lampe, F. C. et al.. Is the prevalence of coronary heart disease falling in British men? Heart 86, 499–505 (2001).

52. Boyle, J. P. et al.. Projection of Diabetes Burden Through 2050: Impact of changing demography and disease prevalence in the U.S. Diabetes Care 24, 1936–1940 (2001).

53. Boulet, L. P. Influence of comorbid conditions on asthma. European Respiratory Journal vol. 33 897–906 (2009).

54. Bardin, P. G., Rangaswamy, J. & Yo, S. W. Managing comorbid conditions in severe asthma. Med. J. Aust. 209, S11-S17.e3 (2018).

55. Lago, R. M., Singh, P. P. & Nesto, R. W. Diabetes and hypertension. Nature Clinical Practice Endocrinology and Metabolism vol. 3 667 (2007).

56. Long, A. N. & Dagogo-Jack, S. Comorbidities of Diabetes and Hypertension: Mechanisms and Approach to Target Organ Protection. Journal of Clinical Hypertension vol. 13 244–251 (2011).

57. de Boer, I. H. et al.. Diabetes and Hypertension: A Position Statement by the American Diabetes Association. Diabetes Care 40, 1273–1284 (2017).

58. Centers for Disease Control and Prevention. Coronary Artery Disease. https://www.cdc.gov/heartdisease/coronary_ad.htm (2019).

59. Rana, J. S., Nieuwdorp, M., Jukema, J. W. & Kastelein, J. J. P. Cardiovascular metabolic syndrome - An interplay of, obesity, inflammation, diabetes and coronary heart disease. Diabetes, Obesity and Metabolism vol. 9 218–232 (2007).

60. Maselli, D. J. & Hanania, N. A. Asthma COPD overlap: Impact of associated comorbidities. Pulmonary Pharmacology and Therapeutics vol. 52 27–31 (2018).

61. Déruaz-Luyet, A. et al.. Multimorbidity and patterns of chronic conditions in a primary care population in Switzerland: A cross-sectional study. BMJ Open 7, e013664 (2017).

62. Cherney, D. Z. I. et al.. Impact of Cardio-Renal-Metabolic Comorbidities on Cardiovascular Outcomes and Mortality in Type 2 Diabetes Mellitus. Am. J. Nephrol. 51, 74–82 (2020).

63. Arnold, S. V. et al.. Burden of cardio-renal-metabolic conditions in adults with type 2 diabetes within the Diabetes Collaborative Registry. Diabetes, Obes. Metab. 20, 2000–2003 (2018).

64. Arnold, S. V. et al.. Cardiovascular Outcomes and Mortality in Type 2 Diabetes with Associated Cardio-Renal-Metabolic Comorbidities. Diabetes 67, 1582–P (2018).

65. Lee, S. J., Lindquist, K., Segal, M. R. & Covinsky, K. E. Development and Validation of a Prognostic Index for 4-Year Mortality in Older Adults. JAMA 295, 808 (2006).

66. Walter, L. C. et al.. Development and Validation of a Prognostic Index for 1-Year Mortality in Older Adults After Hospitalization. JAMA 285, 2987 (2001).

67. Fortin, M., Hudon, C., Haggerty, J., Akker, M. Van Den & Almirall, J. Prevalence estimates of multimorbidity: A comparative study of two sources. BMC Health Serv. Res. 10, 111 (2010).

68. Fortin, M., Stewart, M., Poitras, M.-E., Almirall, J. & Maddocks, H. A Systematic Review of Prevalence Studies on Multimorbidity: Toward a More Uniform Methodology. Ann. Fam. Med. 10, 142–151 (2012).

69. Diederichs, C., Berger, K. & Bartels, D. B. The measurement of multiple chronic diseases - A systematic review on existing multimorbidity indices. Journals Gerontol. - Ser. A Biol. Sci. Med. Sci. 66, 301–311 (2011).

