## Appendix for "Patterns of Multimorbidity"

### Patterns of Multimorbidity (Appendix)

[1]Laboratory for Financial Engineering, Sloan School of Management, Massachusetts Institute of Technology, Cambridge, Massachusetts, United States of America; [2]Department of Electrical Engineering and Computer Science, Massachusetts Institute of Technology, Cambridge, Massachusetts, United States of America; [3]Digital Catalyst, Swiss Re, Cambridge, Massachusetts, United States of America; [4]Sante Fe Institute, Santa Fe, New Mexico, United States of America

**This version: 8 May 2021**

### A Definitions

**Table 1. Age groups based on MeSH definitions.**

| Group | Age |
|---|---|
| Infant | 0 to 2 years |
| Child | 2 to 13 years |
| Adolescent | 13 to 19 years |
| Adult | 19 to 45 years |
| Middle-Aged | 45 to 65 years |
| Aged | 65 to 80 years |
| Elderly | 80 years and over |

**Table 2. List of morbidities and MedDRA System Organ Classes.**

| No. | Disease | MedDRA | Abbrv. |
|---|---|---|---|
| 1 | Angina | Cardiac Disorders | Card |
| 2 | Atrial Fibrillation | Cardiac Disorders | Card |
| 3 | Coronary Artery Disease | Cardiac Disorders | Card |
| 4 | Cardiac Arrhythmia | Cardiac Disorders | Card |
| 5 | Heart Failure | Cardiac Disorders | Card |
| 6 | Heart Valve Disorder | Cardiac Disorders | Card |
| 7 | Myocardial Infarction | Cardiac Disorders | Card |
| 8 | Cardiac-related | Cardiac Disorders | Card |
| 9 | Thyroid Cancer | Endocrine Disorders | En |
| 10 | Osophageal Cancer | Gastrointestinal Disorders | GI |
| 11 | Stomach Cancer | Gastrointestinal Disorders | GI |
| 12 | Liver Disease | Hepatobiliary Disorders | Hep |
| 13 | Liver-related | Hepatobiliary Disorders | Hep |
| 14 | Asthma | Immune System Disorders | Imm |
| 15 | HIV/AIDS | Immune System Disorders | Imm |
| 16 | Multiple Sclerosis | Immune System Disorders | Imm |
| 17 | Diabetes | Metabolism and Nutrition Disorders | Me |
| 18 | Lupus | Musculoskeletal and Connective Tissue Disorders | Mu |
| 19 | Breast Cancer | Neoplasms Benign, Malignant and Unspecified | Neop |
| 20 | Cervix Cancer | Neoplasms Benign, Malignant and Unspecified | Neop |
| 21 | Colorectal Cancer | Neoplasms Benign, Malignant and Unspecified | Neop |
| 22 | Leukemias | Neoplasms Benign, Malignant and Unspecified | Neop |
| 23 | Liver Cancer | Neoplasms Benign, Malignant and Unspecified | Neop |
| 24 | Lung Cancer | Neoplasms Benign, Malignant and Unspecified | Neop |
| 25 | Melanoma | Neoplasms Benign, Malignant and Unspecified | Neop |

| | | | |
|---|---|---|---|
| 26 | Cancer-related | Neoplasms Benign, Malignant and Unspecified | Neop |
| 27 | Pancreas Cancer | Neoplasms Benign, Malignant and Unspecified | Neop |
| 28 | Encephalitis | Nervous System Disorders | Nerv |
| 29 | Parkinson's Disease | Nervous System Disorders | Nerv |
| 30 | Stroke | Nervous System Disorders | Nerv |
| 31 | Stroke Related | Nervous System Disorders | Nerv |
| 32 | Dementia | Psychiatric Disorders | Ps |
| 33 | Bladder Cancer | Renal and Urinary Disorders | Ren |
| 34 | Kidney Disease | Renal and Urinary Disorders | Ren |
| 35 | Kidney-related | Renal and Urinary Disorders | Ren |
| 36 | Ovarian Cancer | Reproductive System and Breast Disorders | Repr |
| 37 | Prostate Cancer | Reproductive System and Breast Disorders | Repr |
| 38 | Testicular Cancer | Reproductive System and Breast Disorders | Repr |
| 39 | Uterus Cancer | Reproductive System and Breast Disorders | Repr |
| 40 | Chronic Obstructive Pulmonary Disease | Respiratory, Thoracic and Mediastinal Disorders | Res |
| 41 | Respiratory-related | Respiratory, Thoracic and Mediastinal Disorders | Res |
| 42 | Aortic Aneurysm | Vascular Disorders | Vas |
| 43 | Hypertension | Vascular Disorders | Vas |
| 44 | Other Aneurysms | Vascular Disorders | Vas |
| 45 | Peripheral Artery Disease | Vascular Disorders | Vas |
| 46 | Transient Ischemic Attack | Vascular Disorders | Vas |

### B Sample Size for Index of Multiple Deprivation

| **Millions** | **2008** | **2009** | **2010** | **2011** | **2012** | **2013** | **2014** | **2015** | **2016** |
|---|---|---|---|---|---|---|---|---|---|
| Total | 5.1 | 5.1 | 5.0 | 5.0 | 4.9 | 4.6 | 4.2 | 3.5 | 3.1 |
| With IMD | 0.3 | 2.1 | 2.2 | 2.1 | 2.1 | 1.9 | 1.8 | 1.4 | 1.2 |
| *% of Total* | *5.2* | *42.0* | *43.4* | *41.3* | *43.4* | *41.6* | *42.0* | *39.8* | *37.2* |

### C Supplementary Figures

Supplementary plots for all age groups and years at available at https://www.dropbox.com/sh/iljxpdll783zy1m/AAAIFy2fHIW0N40Zvzv7srj7a?dl=0.

### D Survival Analysis

**Table 3. Test set C-index of survival models.**

| | Test Set C-index (95% CI) | | | |
|---|---|---|---|---|
| Year | Cox | Regularized Cox | Accelerated Failure Time | Neural Network Survival |
| 2010 | 0.793 (0.790, 0.797) | 0.793 (0.790, 0.797) | 0.793 (0.790, 0.797) | 0.794 (0.790, 0.797) |
| 2011 | 0.798 (0.795, 0.802) | 0.798 (0.795, 0.802) | 0.798 (0.795, 0.802) | 0.798 (0.794, 0.801) |
| 2012 | 0.807 (0.803, 0.811) | 0.807 (0.804, 0.812) | 0.799 (0.795, 0.804) | 0.806 (0.802, 0.810) |
